# Accelerated aortic stiffness is associated with brain structure, perfusion and cognition in the Whitehall II Imaging Sub-study

**DOI:** 10.1101/2020.07.01.20142612

**Authors:** Sana Suri, Scott T. Chiesa, Enikő Zsoldos, Clare E. Mackay, Nicola Filippini, Ludovica Griffanti, Abda Mahmood, Archana Singh-Manoux, Martin J Shipley, Eric J Brunner, Mika Kivimäki, John E. Deanfield, Klaus P. Ebmeier

**Affiliations:** Department of Psychiatry, Warneford Hospital, University of Oxford, OX3 7JX, Oxford, United Kingdom; Wellcome Centre for Integrative Neuroimaging, University of Oxford, OX3 7JX, Oxford, United Kingdom; Institute of Cardiovascular Science, University College London, EC1A 4NP, London, United Kingdom; Department of Epidemiology and Public Health, University College London, WC1E 6BT London, United Kingdom; Inserm U1153, Epidemiology of Ageing and Neurodegenerative diseases, Université de Paris, 75010 Paris, France

**Author notes:** Joint first authorship (Sana Suri, Scott T. Chiesa). Joint last authorship (John E. Deanfield, Klaus P. Ebmeier). Corresponding author: Dr Sana Suri.

**Keywords:** aortic stiffness, pulse wave velocity, ageing, brain magnetic resonance imaging, diffusion imaging, cerebral perfusion

## Abstract

**Background:** Aortic stiffness is closely linked with cardiovascular diseases, but recent studies suggest that it is also a risk factor for cognitive decline and dementia. However, the brain changes underlying this risk are unclear. We examined whether aortic stiffening in the transition from mid to late-life affects brain structure and cognition.

**Methods and Findings:** Aortic pulse wave velocity was measured in 2007-09 (Phase 9) and at a 4-year follow-up in 2012-13 (Phase 11) in the Whitehall II Imaging Sub-study cohort. Between 2012-2016 (Imaging Phase), participants received a multi-modal 3T brain magnetic resonance imaging (MRI) scans and cognitive tests. Participants were selected if they had no clinical diagnosis of dementia and no gross brain structural abnormalities. Voxel-based analyses were used to assess grey matter volume, white matter microstructure (fractional anisotropy and diffusivity), cerebral blood flow, and white matter lesions. Cognitive outcomes were performance on verbal memory, semantic fluency, working memory and executive function tests. Of 544 participants, 445 (81.8%) were men. The mean (SD) age was 63.9 (5.2) years at the baseline Phase 9 examination, 67.9 (5.3) years at Phase 11 and 69.8 (5.2) years at the Imaging Phase. Voxel-based analysis revealed that accelerated aortic stiffening in mid-to-late life was associated with poor white matter integrity, viz. lower fractional anisotropy in 4.2% of white matter and higher radial diffusivity in 6.7% of white matter, including the corpus callosum, corona radiata, superior longitudinal fasciculus and corticospinal tracts. Accelerated aortic stiffening was also related to lower cerebral perfusion in 1.1% of grey matter including the parietal, frontal, and occipital cortices. No associations with grey matter volume or white matter lesions were observed. Further, higher baseline aortic stiffness was associated with poor semantic fluency (B=-0.48, 95%CI −0.77 to −0.19, p<0.005) and verbal learning outcomes (B=-0.36, 95%CI - 0.60 to −0.13, p<0.005).

**Conclusions:** Faster aortic stiffening in mid-to-late life is associated with poor brain white matter microstructural integrity and reduced cerebral perfusion, likely due to increased transmission of pulsatile energy to the delicate cerebral microvasculature. Strategies to prevent arterial stiffening prior to this point may be required to offer cognitive benefit in older age.

## Introduction

Nearly 50 million people currently live with dementia worldwide and this figure is expected to triple by 2050.[1] While the exact mechanisms underlying dementia are still poorly understood, accumulating evidence implicates a number of potentially modifiable risk factors linked to another global health burden, cardiovascular disease (CVD).[2] Timely prevention strategies targeting risk factors common to these diseases may therefore have a dual benefit in reducing two of the world’s most prevalent causes of morbidity and mortality.

One established risk factor for CVD is disproportionate stiffening of the aorta.[3] Aortic stiffness refers to the loss of elasticity in the artery wall and occurs gradually with age.[4] Acceleration of this process can also compromise the artery’s ability to dampen the pulsatile energy flowing from the heart to delicate target organs like the brain. Indeed, numerous studies have now demonstrated a relationship between aortic stiffness and accelerated cognitive decline, suggesting that this may also be an important risk factor for dementia ([5–8], for review see[9]). Less well understood, however, is the relationship of aortic stiffness with cerebrovascular and microstructural changes in the brain that may contribute to cognitive impairment and dementia.[10]

Some magnetic resonance imaging (MRI) studies have revealed a link between higher aortic stiffness and markers of vascular pathology in the brain such as white matter lesions (WMLs), lacunar infarcts, and microbleeds ([11,12], for review see[13]). However, most of these studies have visually dichotomized brain tissue into normal and abnormal, which lacks quantitative and spatial detail about the specific brain regions that may be most sensitive to damage. Very few studies have examined brain microstructural[14–16] or cortical perfusion[17] correlates of aortic stiffness at a voxel level.

Moreover, to our knowledge, no human study to date has investigated the potential impact that accelerated rates of arterial stiffening have on these measures. Arterial stiffening is commonly observed in the transition from mid to late life; a time when a combination of ageing and long-term CVD risk exposure interact to negatively influence distensibility of major arteries. Recent work in animal models has strengthened claims that this link between excess pulsatility and cerebral outcomes may be causal.[18] De Montgolfier *et al*. showed that surgically induced rapid increases in pulsatile pressure in mice results in endothelial dysfunction and hypoperfusion within the brain’s fragile microcirculation, all accompanied by significant declines in cognitive function.[18]

Using repeat measures of carotid-femoral pulse wave velocity (PWV) measured as part of the long-running Whitehall II study, we assessed changes in arterial stiffness over 4 years, and investigated their relationship with multi-modal brain MRI and cognitive outcomes in a subset of 544 randomly-selected participants in the Whitehall II Imaging Sub-Study.

## Methods

### Sample selection

Participants were drawn from the Whitehall II study, a cohort of 10,308 British Civil Servants followed since 1985.[19] Arterial stiffness was measured using carotid-femoral pulse wave velocity (PWV), at Phase 9 (2007-2009, n=4347) and again in Phase 11 (2012-2013, n=4344).[20] Following Phase 11, 775 randomly selected participants received multi-modal brain MRI scans and cognitive tests at the Wellcome Centre for Integrative Neuroimaging, University of Oxford, as part of the Whitehall II Imaging Sub-study (2012-2016, protocol described previously[21]). Sample selection is in S1 Figure. The study was approved by the University of Oxford Medical Sciences Interdivisional Research Ethics Committee and written informed consent was obtained from all participants.

### Pulse Wave Velocity

The protocol for measuring PWV in the Whitehall II study has been described previously.[6] Briefly, aortic PWV was assessed between carotid and femoral sites using applanation tonometry (SphygmoCor; Atcor Medical, Australia). At each phase, two measurements of PWV were acquired from each participant, and a third measurement was taken if the difference between the first two measurements was >0.5 m/s. The average of the measurements at each phase was used in cross-sectional analyses. The change (ΔPWV) was calculated as the difference between phases divided by time between visits (m/s/year), with positive values representing faster stiffening.

### Neuroimaging outcomes

Detailed protocols for the acquisition and pre-processing of all scans have been described previously[21] and in the Supplementary Information (S3 Protocol). All images were processed using FMRIB Software Library (FSL) tools. Grey matter (GM) volumes were assessed using voxel-based morphometry analyses (FSL-VBM) on structural T1-weighted scans.[22] White matter lesions (WMLs) were quantified as hyperintensities on fluid-attenuated inversion recovery scans using the FSL-BIANCA tool.[23] Cerebral blood flow (CBF) was quantified as the rate of delivery of arterial blood to brain tissue (ml of blood per 100g of tissue per minute) using pseudocontinous arterial spin labelling analysed with the FSL-BASIL toolkit.[24] White matter microstructure was assessed using diffusion tensor imaging (DTI) scans analysed with tract-based spatial statistics (FSL-TBSS).[25] DTI is sensitive to directional diffusion of water within the axon, which is unrestricted along the axon, but hindered perpendicularly due to the presence of the myelin sheath. Directionality and amount of water diffusion is quantified by DTI-derived metrics such as fractional anisotropy (FA), radial diffusivity (RD), axial diffusivity (AD) and mean diffusivity (MD).

Decreases in FA alongside increases in diffusivity are established and sensitive measures of WM damage in dementia.[26] Lower CBF and higher WMLs are also well-established surrogate neuroimaging end points for dementia.[27]

In addition to voxel-based analyses, we also examined summary measures of global brain physiology. For each subject, total GM and WML volumes were extracted using FSL-FAST and FSL-BIANCA, respectively. These were normalised to total brain volume and expressed as GM% and WML%. Total FA and diffusivity metrics were extracted from the mean TBSS skeleton, and total CBF from the frontal, temporal, parietal and occipital lobes were extracted from native-space CBF maps using MNI152 masks.

Due to a scanner upgrade two-thirds of the way through the study, two MRI scanners were used: a 3T Siemens Magnetom Verio scanner with a 32-channel head coil (n=552, April 2012–December 2014) and a 3T Siemens Prisma Scanner with a 64-channel head-neck coil in the same centre (n=222, July 2015–December 2016).[21] The scan parameters were identical or closely matched between scanners, and a scanner covariate was used in all analyses. CBF scans were acquired in a subset of 145 participants who were all scanned on the Verio scanner.

### Cognitive outcomes

We examined performance on seven cognitive domains; *semantic fluency*: number of animals named in one minute; *verbal learning:* number of words learned after 3 trials of the Hopkins Verbal Learning Test (HVLT-R), *delayed verbal recall*: number of words recalled after a 30-minute delay on HVLT-R; *working memory*: total score on the Digit Span Forward, Backward and Sequence tests; *executive function*: difference in time between Trail Making Test B and A, and digit substitution test (WAIS-IV); *global cognitive function:* Montreal Cognitive Assessment (MoCA).

### Covariates

Covariates were selected *a priori* based on the literature, and included age, mean arterial pressure (MAP), body mass index (BMI) and anti-hypertensive treatment each reported at the respective phase (9 or 11), sex, total years of full-time education (reported at the MRI visit), socioeconomic status (grades 1-3 reported at Phase 7 in 2002-2004), number of years between repeat visits, and MRI scanner model. Additional covariates such as current smoking status (yes/no), current cardiovascular disease (yes/no), history of type 2 diabetes (yes/no) were tested in a stepwise forward linear regression and as they did not significantly improve model fit they were not included in the final results described below to avoid overfitting.

### Statistical analysis

Longitudinal change in PWV was analysed using linear mixed effects regression using the nlme package in R, with a continuous autoregressive moving-average correlation structure to consider correlations between repeated measures on the same individual. The intercepts and slopes were fitted as random effects.

Cross-subject voxel-wise multivariable regressions were performed separately on spatial maps of GM volume, white matter microstructure (FA, MD, RD and AD) and CBF (pre-processing detailed in the supplementary file, S3 Protocol). For each modality, the respective spatial maps for all subjects were concatenated into a 4D file. This was submitted to FSL-Randomise to perform a permutation-based nonparametric test with 5000 permutations, using the threshold-free cluster enhancement option. Three general linear models (GLM) were run to assess the associations of Phase 9 PWV (Model 1), ΔPWV (Model 2) and Phase 11 PWV (Model 3) on the aforementioned MRI and cognitive outcomes. All models were adjusted for covariates, and voxel-wise results are displayed at a significance threshold of p<0.05 corrected for multiple comparisons across space.

Statistical analyses of MRI-derived variables were performed in R (Version 1.1.463). WML% and GM%, were used as dependent variables in multivariable linear regressions against Phase 9 (Model 1), ΔPWV (Model 2), and Phase 11 PWV (Model 3). Lobar CBF (frontal, temporal, parietal, occipital), WM metrics (global FA, MD, RD and AD), and the seven cognitive tests were used as dependent variables in separate multivariate GLMs to test for associations with Phase 9 PWV (Model 1), ΔPWV (Model 2), and Phase 11 (Model 3). Post-hoc univariate regressions were performed to assess contributions of the individual CBF, WM and cognitive measures. We assessed whether associations of PWV with cognition were mediated by brain MRI markers using a mediation analysis in PROCESSv3.4 for SPSS (2012-2019 by Andrew F. Hayes; data not shown). We also examined the association of brain markers with cognitive performance using partial correlations adjusted for age, sex, years of education, scanner type, and socioeconomic status.

## Results

### Participant characteristics

Participant characteristics are in Tables 1 and 2; 81.8% of the participants were male, and 58.6%, 38.4% and 2.9% of participants were categorised in socioeconomic grades 1, 2 and 3 respectively. There was a significant increase in PWV with time after adjusting for baseline age, sex and socioeconomic status (B=0.17, 95%CI 0.14 to 0.20, p<0.0001). On average, PWV increased by 0.2(0.4) m/s per year (S2 Figure).

**Table 1:** Characteristics of the 544 participants at Phase 9 and Phase 11. Values represent mean (standard deviation) or No. (%).

| **Sample Characteristics** | **Phase 9**<br>**2007-2009** | **Phase 11**<br>**2012-2013** |
| --- | --- | --- |
| Age, years | 63.9 (5.2) | 68.0 (5.3) |
| Age range, years | 55.6-77.4 | 59.8–81.8 |
| Sex, %male (N) | 81.8% (445/544) |  |
| Pulse wave velocity m/s | 8.1 (1.7) | 8.8 (2.2) |
| Pulse wave velocity range m/s | 4.7 -15.0 | 4.5 - 20.0 |
| Systolic blood pressure, mmHg | 122.5 (14.7) | 125.2 (15.6) |
| Diastolic blood pressure, mmHg | 70.6 (10.1) | 70.4 (9.5) |
| Mean arterial pressure, mmHg | 87.9 (11.0) | 88.6 (10.7) |
| Body Mass Index, kg/m<sup>2</sup> | 25.9 (3.6) | 25.9 (3.6) |
| Anti-hypertensive treatment, No. (%) | 139/544 (25.6%) | 177/544 (32.5%) |
| Current smokers, No. (%) | 29/540 (5.3%) | 20/543 (3.7%) |
| Type II diabetes, No. (%) | 8/544 (1.5%) | 38/541 (7.0%) |
| Current CVD (angina, stroke or myocardial infarction), No. (%) | 12/544 (2.2%) | 16/544 (2.9%) |

**Table 2:** Summary of the brain and cognitive outcome variables examined at the MRI Phase (2012-2016).

| OVERVIEW | Mean (SD) |
| --- | --- |
| Age at MRI Phase, years | 69.82 (5.23) |
| Years of education | 14.23 (3.01) |
| Years from Phase 9 to MRI Phase | 5.90 (1.39) |
| Years from Phase 11 to MRI Phase | 1.82 (1.85) |
| Years from Phase 9 to Phase 11 | 4.08 (0.18) |
| MRI OUTCOMES |  |
| Grey matter volume, % TBV | 38.40 (1.87) |
| Global fractional anisotropy ($\times 10^{-1}$) | 4.80 (0.20) |
| Global mean diffusivity ($\times 10^{-4}$) | 6.85 (0.28) |
| Global radial diffusivity ($\times 10^{-4}$) | 4.86 (0.30) |
| Global axial diffusivity ($\times 10^{-4}$) | 10.82 (0.26) |
| White matter lesions, % TBV<sup>a</sup> | 0.40 (0.26) |
| Frontal Lobe CBF (ml/100g/min)<sup>b</sup> | 51.52 (12.22) |
| Parietal Lobe CBF (ml/100g/min)<sup>b</sup> | 62.15 (15.44) |
| Occipital Lobe CBF (ml/100g/min)<sup>b</sup> | 49.04 (18.74) |
| Temporal Lobe CBF (ml/100g/min)<sup>b</sup> | 49.48 (10.21) |
| COGNITIVE OUTCOMES<sup>c</sup> |  |
| Verbal learning (short-term recall) | 27.80 (4.37) |
| Delayed verbal recall | 9.45 (2.51) |
| Semantic Fluency | 22.49 (5.28) |
| Short term memory (Total Digit Span) | 31.06 (5.47) |
| Executive Function (Trail Making Test B-A) | 0.58 (0.44) |
| Executive Function (Digit Substitution) | 63.41 (13.05) |
| Montreal Cognitive Assessment | 27.38 (2.11) |

Values represent mean (standard deviations). Grey matter volume and white matter lesions are expressed as % of total brain volume, TBV. Abbreviations: CBF, cerebral blood flow.
<sup>a</sup>Analysed white matter lesion subset (N=535)
<sup>b</sup>Analysed CBF subset (N=113)
<sup>c</sup>Analysed cognitive subset (N=539)

### Association of PWV with brain microstructure

Higher ΔPWV was the strongest predictor of poor WM microstructure in covariate-adjusted multivariate GLMs with global FA, MD, RD, AD as dependent variables (p_(model)_=0.015, summary statistics in Table 3). Post-hoc linear regressions revealed that this effect was driven by associations of aortic stiffening with lower FA and higher RD (Table 3). Voxel-wise analyses demonstrated that associations with FA and RD were localised to 4.2% and 6.7% of white matter tracts respectively, covering the corpus callosum, fornix, left anterior thalamic radiation, superior corona radiata, and corticospinal tracts. Associations with RD were further distributed within the superior and inferior longitudinal fasciculus, posterior corona radiata, and bilateral external and internal capsule (Figure 1). While evidence of associations were also seen with *cross-sectional* PWV, these were less pronounced both with the extracted WM measures [Phase 11 PWV: (p_(model)_ =0.019); Phase 9 PWV: (p_(model_ =0.29) Table 3] and voxel-based analyses (data not shown). Neither cross-sectional nor longitudinal aortic PWV were associated with WML%, total GM% (Table 3) or GM volume assessed with voxel-wise analysis.

**Table 3:** Associations between PWV and brain and cognitive outcomes.

| Outcome<sup>1</sup> | Model 1: Phase 9 PWV (m/s) | | Model 2: ΔPWV (m/s/y) | | Model 3: Phase 11 (m/s) | |
| --- | --- | --- | --- | --- | --- | --- |
|  | B [95% CI] | p | B [95% CI] | p | B [95% CI] | p |
| **Brain MRI Outcomes** |  |  |  |  |  |  |
| Fractional Anisotropy<sup>2</sup> | -0.66 [-1.65, 0.32] | 0.19 | -4.16 [-8.19, -0.13] | **0.04** | -0.73 [-1.54, 0.08] | 0.08 |
| Mean Diffusivity<sup>2</sup> | 0.91 [-0.45, 2.28] | 0.19 | 4.05 [-1.55, 9.64] | 0.16 | 0.79 [-0.33, 1.91] | 0.17 |
| Radial Diffusivity<sup>2</sup> | 1.06 [-0.41, 2.53] | 0.16 | 5.54 [-0.47, 11.55] | 0.07 | 1.05 [-0.16, 2.25] | 0.09 |
| Axial Diffusivity<sup>2</sup> | 0.61 [-0.70, 1.92] | 0.36 | 1.06 [-4.32, 6.44] | 0.70 | 0.28 [-0.79, 1.36] | 0.61 |
| Grey Matter % | -0.03 [-0.12, 0.06] | 0.54 | -0.06 [-0.44, 0.32] | 0.75 | -0.04 [-0.11, 0.04] | 0.36 |
| White Matter Lesions %<sup>3</sup> | -0.003 [-0.02, 0.01] | 0.71 | 0.006 [-0.05, 0.06] | 0.84 | -0.003 [-0.01, 0.01] | 0.60 |
| Frontal CBF<sup>4</sup> | 0.75 [-0.98, 2.5] | 0.39 | -6.99 [-13.64, -0.35] | **0.04** | -0.49 [-1.78, 0.80] | 0.45 |
| Temporal CBF<sup>4</sup> | 0.74 [-0.66, 2.13] | 0.30 | -4.37 [-9.78, 1.05] | 0.11 | -0.02 [-1.08, 1.04] | 0.98 |
| Parietal CBF<sup>4</sup> | 0.64 [-1.50, 2.78] | 0.56 | -8.91 [-17.12, -0.69] | **0.03** | -0.75 [-2.4, 0.85] | 0.35 |
| Occipital CBF<sup>4</sup> | 0.69 [-1.84, 3.21] | 0.59 | -10.16 [-19.88, -0.44] | **0.04** | -1.04 [-2.91, 0.82] | 0.27 |
| **Cognitive Outcomes<sup>5</sup>** |  |  |  |  |  |  |
| Semantic Fluency | -0.48 [-0.77, -0.19] | **0.001** | 0.81 [-0.36, 1.97] | 0.18 | -0.14 [-0.38, 0.10] | 0.24 |
| Verbal Learning | -0.36 [-0.60, -0.13] | **0.003** | 0.43 [-0.54, 1.39] | 0.39 | -0.10 [-0.30, 0.09] | 0.29 |
| Delayed Verbal Recall | -0.11 [-0.25, 0.03] | 0.12 | 0.20 [-0.36, 0.77] | 0.48 | -0.003 [-0.12, 0.11] | 0.97 |
| Working Memory (Digit Span) | -0.03 [-0.33, 0.27] | 0.86 | -0.17 [-1.40, 1.07] | 0.79 | -0.08 [-0.33, 0.17] | 0.53 |
| Executive Function (Trail Making) | -0.003 [-0.03, 0.02] | 0.79 | -0.0 [-0.12, 0.07] | 0.62 | -0.005 [-0.03, 0.02] | 0.62 |
| Executive Function (Digit Substitution) | -0.33 [-1.03, 0.37] | 0.36 | 1.41 [-1.46, 4.28] | 0.34 | -0.08 [-0.66, 0.50] | 0.79 |
| Global Cognition (MOCA) | -0.14 [-0.26, -0.02] | **0.02** | -0.15 [-0.62, 0.32] | 0.54 | -0.09 [-0.19, 0.006] | 0.07 |

<sup>1</sup>Aforementioned covariates were used in each model. In Model 2, Phase 9 PWV and years from P11 to MRI were also added to examine the unique contribution of rate of stiffening (regardless of baseline).
<sup>2</sup>FA: B [95% CI] x 10<sup>-3</sup> and MD, RD, AD: B [ 95% CI] x 10<sup>-6</sup>
<sup>3</sup>Analysed white matter lesions subset, N=535
<sup>4</sup>Analysed CBF subset, N=113
<sup>5</sup>Analysed cognitive subset, N=539

**Figure 1:**
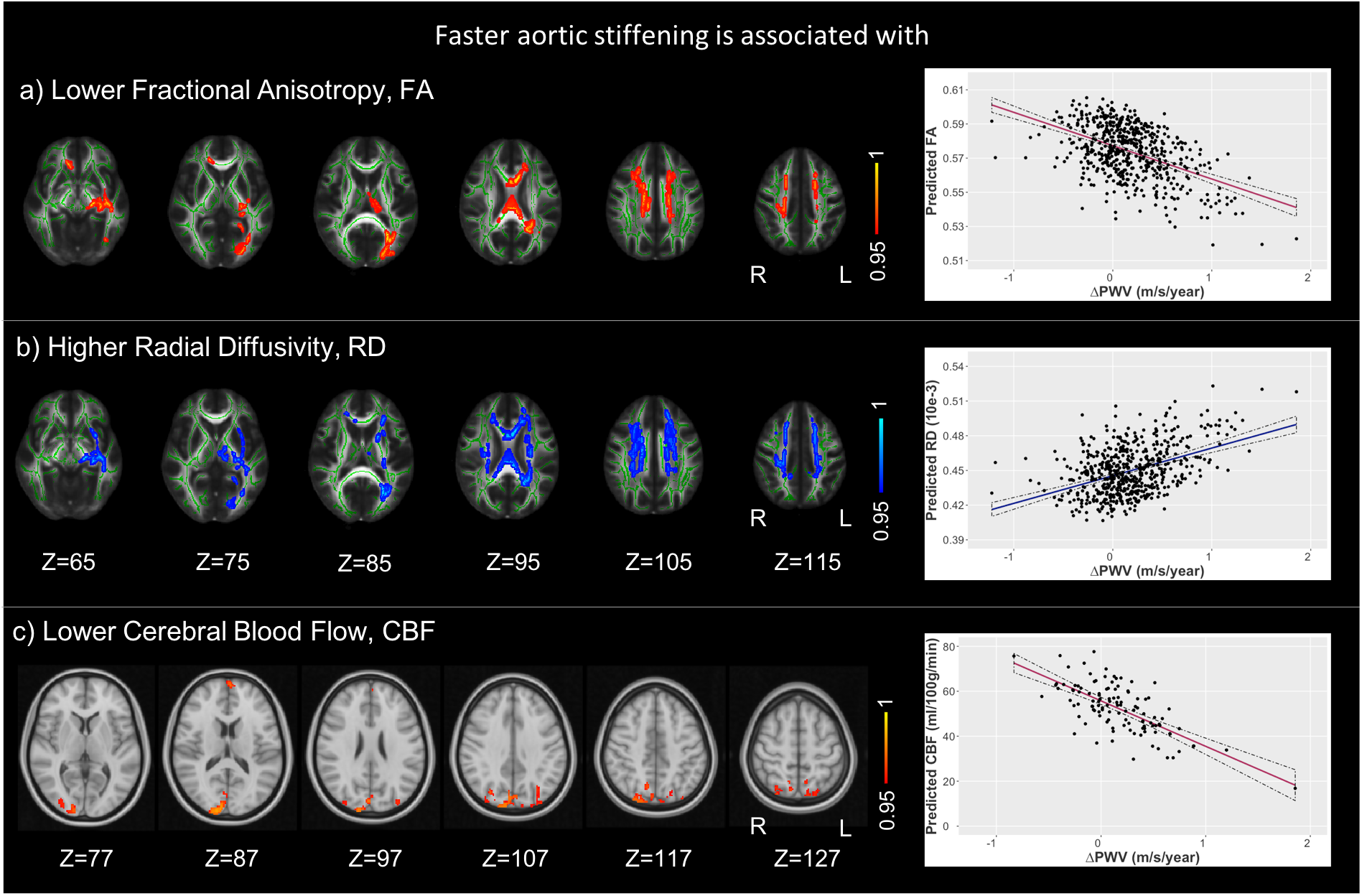
Association of rate of aortic stiffening white matter microstructure and cerebral blood flow. Faster aortic stiffening was associated with (A) lower fractional anisotropy (FA), (B) higher radial diffusivity (RD), and (C) lower cerebral blood flow (CBF). Six horizontal slices are displayed ranging with MNI coordinates ranging from Z=65 to Z=115 for FA, RD and Z=77 to Z=127 for CBF. The FA and RD clusters are overlaid on the study-specific mean FA skeleton (green) and the standard FMRIB58 FA image, and the CBF clusters are overlaid on the standard MNI152 brain. All results are thresholded at TFCE-corrected p<0.05 and are presented with a colour gradient for 1-p values. Scatter plots represent the estimated marginal means for predicted FA, RD and CBF extracted from the significant clusters, and are adjusted for covariates. Abbreviations: R, right; L, left

### Association of PWV with cerebral perfusion

Higher ΔPWV was associated with lower total CBF in the frontal, parietal and occipital lobes (Table 3). The voxel-wise analyses revealed most pronounced effects in the parietal, lateral occipital and cuneal cortex, occipital pole, frontal pole, and parts of the precuneus (Figure 1). *Cross-sectional* PWV at Phases 9 and 11 were not significantly associated with regional CBF in the voxel-wise analysis or using extracted mean lobar CBF (Table 3).

### Association of PWV with cognitive performance

Baseline PWV at Phase 9 was associated with lower overall cognitive performance at the MRI phase (p_(model)_=0.004) in covariate-adjusted multivariate GLMs. Post-hoc linear regressions revealed that this association was driven by poorer performance on the semantic fluency and verbal learning tests (Table 3). In contrast, neither Phase 11 PWV (p_(model)_=0.37) nor the rate of aortic stiffening (p_(model)_=0.70) were associated with cognitive performance at the MRI visit (Table 3).

### Association of cerebral measures and cognitive performance

WM microstructure and CBF were associated with executive function and working memory (Table 4). However, these brain measures did not mediate the aforementioned relationship between baseline PWV and semantic fluency and verbal learning.

**Table 4:** Partial correlations of cognitive function with PWV-associated brain MRI outcomes, correcting for age, sex, years of education, socio-economic status and scanner model.

|  | **Fractional Anisotropy<sup>1</sup>**<br>**(r, p)** | **Mean Diffusivity<sup>1</sup>**<br>**(r, p)** | **Radial Diffusivity<sup>1</sup>**<br>**(r, p)** | **Total Cerebral Blood Flow<sup>2</sup>**<br>**(r, p)** |
| --- | --- | --- | --- | --- |
| **Semantic Fluency** | 0.09, 0.05 | -0.05, 0.23 | -0.07, 0.13 | -0.07, 0.46 |
| **Verbal Learning** | 0.08, 0.07 | -0.05, 0.26 | -0.06, 0.17 | -0.02, 0.84 |
| **Delayed Verbal Recall** | 0.08, 0.06 | -0.06, 0.18 | -0.07, 0.10 | 0.01, 0.91 |
| **Working Memory (Digit Span)** | **0.13, 0.002** | **-0.11, 0.01** | **-0.12, 0.006** | 0.11, 0.27 |
| **Executive Function (Trail Making)** | -0.09, 0.04 | 0.07, 0.11 | 0.08, 0.07 | **-0.25, 0.008** |
| **Executive Function (Digit Substitution)** | **0.14, 0.001** | **-0.11, 0.01** | **-0.13, 0.003** | 0.15, 0.13 |
| **Global Cognition (MOCA)** | 0.08, 0.07 | -0.06, 0.20 | -0.07, 0.13 | 0.18, 0.07 |

<sup>1</sup>Analysed cognitive subset (N=539)
<sup>2</sup>Analysed CBF subset (N=113)

## Discussion

In one of the largest studies to-date relating mid-life PWV changes to extensive cerebral phenotyping and cognitive outcomes, we show that an increased rate of arterial stiffening is associated with lower white matter microstructural integrity and cerebral blood flow in older age. Furthermore, these associations were present in diffuse brain areas, suggesting that exposure to excess pulsatility may result in a widespread damaging effect on the fragile cerebral microstructure. Cognitive function at follow-up related more closely with baseline arterial stiffness rather than rate of arterial stiffening. Taken together, these findings suggest that although accelerated arterial stiffening in the transition to old age may negatively impact brain structure and function, long-term exposure to higher levels of arterial stiffness prior to this point may be the most important determinant for future cognitive ability.

Patients with Alzheimer’s disease and vascular dementia have higher levels of aortic stiffness relative to cognitively healthy adults, suggesting that arterial dysfunction may play a role in dementia.[28] Aortic stiffening is a hallmark of vascular ageing and may lead to a heightened state of oxidative and inflammatory damage within the cerebral tissues due to an increased penetrance of excess pulsatility into the fragile microcirculation of the brain.[9] These changes have been shown to disrupt endothelial cell function and the blood brain barrier in animal models, and have also been hypothesised to compromise cerebral perfusion and ultimately lead to neurodegeneration and cognitive impairment.[9] While previous studies have related cross-sectional measures of arterial stiffness to cognition, this is the first study to publish associations between progressive increases in aortic stiffening over a 4-year period and cerebral and cognitive outcomes in later life.

We report a number of novel findings. First, accelerated arterial stiffening was found to associate with WM microstructural decline within the corpus callosum, corona radiata, superior longitudinal fasciculus, cortico-spinal tracts and internal capsule. Observations of lower FA with a concomitantly higher RD within these tracts are suggestive of axonal degeneration and loss of myelin. Notably, these tracts have recently been shown to be vulnerable to arterial stiffness in younger adults (∼50 years old) from the Framingham Heart Study[15], as well as two smaller studies in older adults.[14,16] These tracts are supplied by the anterior and middle cerebral arteries and given that they are also compromised in Alzheimer’s and vascular dementia[26] they may be potential markers of the role of arterial stiffening in dementia pathophysiology. In contrast to some other studies[11,12,15], however, we noted no associations of aortic stiffness with WMLs or GM volumes. It is possible that our cohort – which compared to the general UK population is relatively high-functioning with a healthier vascular profile[19] – had a relatively lower WML burden. Thus, changes in WM diffusion metrics (which have been noted to precede larger volumetric and pathological changes) may have been more sensitive to the underlying structural tissue alterations at a voxel level. Future studies examining longitudinal changes in GM atrophy or WML burden would be more informative than measures from a single time-point.

Second, in addition to evidence of disruptions to WM structural integrity, we also noted significantly lower CBF in posterior cingulate, cuneal, frontal and occipital cortices. These results are in line with a recent cross-sectional study in individuals with mild cognitive impairment[17], and implicate brain areas typically affected by vascular ageing.[29] Similar to WM metrics, these effects were found to be 5-7 times larger for rate of stiffening than cross-sectional PWV, suggesting that exposure to rapid increases in pulsatile stress may be particularly damaging to cerebral perfusion compared with more gradual changes over time.

Third, we observed an inverse association between baseline PWV and semantic fluency and verbal learning. This supports data from this and other cohorts[6–8] which suggest a possible influence of arterial stiffness on these two domains that are preferentially impaired in Alzheimer’s disease.[30] In addition, we found evidence of a weak association between aortic stiffness and poor performance on the MoCA, which is used to screen for amnestic mild cognitive impairment. These cognitive outcomes, however, were associated with baseline aortic stiffness alone, suggesting that long-term exposure to arterial stiffness in early life may be the most important factor determining these cognitive abilities in older age. Thus, interventions to reduce exposure to risk factors earlier in life may be required to in order to offer maximal benefits to later-life cognition. However, our observation of an additional relationship between accelerated arterial stiffening after this point and further detrimental changes to WM structure and cerebral perfusion provides the first evidence of a potential mechanism underlying the continued increase in rate of cognitive decline, which has repeatedly been demonstrated in this and other cohorts during the transition into later life.[6,31]

This study has a number of strengths and limitations. This is one of the largest MRI cohorts with multi-modal brain imaging phenotyping, extensive cognitive testing, and longitudinal arterial stiffness measures. Moreover, arterial stiffness was measured six years prior to the MRI scan, starting at a mean age of 64 years when vascular risk has been demonstrated to contribute to dementia pathology. However, as with all cohort studies, it is not possible to infer causal associations between these exposures and outcomes. In addition, with this relatively well-educated and predominantly male cohort it is possible that we are underestimating the observed association between PWV and cognition. Nonetheless, as we have adjusted for sex and education in all our regression models, it is unlikely that these variables have significantly driven our results. Last, although we examined the role of longitudinal changes in PWV on prospective brain structure, we were limited by cross-sectional MRI measurements.

### Conclusion

We provide the first evidence of a relationship between accelerated aortic stiffening in mid to late-life and detrimental changes to cerebral white matter microstructural integrity and perfusion in later life. These findings suggest that diffusion and perfusion MRI may be sensitive markers of recent damage caused by aortic stiffening and could therefore be potentially relevant outcome measures in intervention studies. However, as measures of amnestic cognition were found to most closely relate to baseline measures of stiffness, such interventions may need to be targeted earlier in life in order to offer maximal benefit for both vascular and cognitive health.

## Data Availability

The study follows Medical Research Council data sharing policies (https://mrc.ukri.org/research/policies-and-guidance-for-researchers/data-sharing/). In accordance with these guidelines, data from the Whitehall II Imaging Sub-study will be accessible via the Dementias Platform UK (https://portal.dementiasplatform.uk/). Data from the Whitehall II Study is available through formal request to the data sharing committee (https://www.ucl.ac.uk/iehc/research/epidemiology-public-health/research/whitehallII/data-sharing).

https://www.ucl.ac.uk/iehc/research/epidemiology-public-health/research/whitehallII/data-sharing

## Funding

The Whitehall II study is supported by the British Heart Foundation (RG/16/11/32334), UK Medical Research Council (K013351) and US National Institute on Aging (R01AG013196; R01AG034454; R01AG056477). The Whitehall II Imaging Sub-study was supported by the UK Medical Research Council (MRC) grants “Predicting MRI abnormalities with longitudinal data of the Whitehall II Sub-study” (G1001354; PI KPE; ClinicalTrials.gov Identifier: NCT03335696), and “Adult Determinants of Late Life Depression, Cognitive Decline and Physical Functioning - The Whitehall II Ageing Study” (MR/K013351/1; PI: MK). Working on this study was also supported by European Commission (Horizon 2020 grant “Lifebrain”, 732592; co-PI KPE), the HDH Wills 1965 Charitable Trust (1117747; PI KPE) and the UK National Institute of Health Research (NIHR) Oxford Health Biomedical Research Centre (BRC). The Wellcome Centre for Integrative Neuroimaging (WIN) is supported by core funding from the Wellcome Trust (203139/Z/16/Z). The authors report the following funding: **SS** (Alzheimer’s Society Research Fellowship (Grant Number 441)), **KPE** (UK Medical Research Council (G1001354, MR/K013351/), HDH Wills 1965 Charitable Trust (1117747), Alzheimer Research UK (PPG2012A-5) and the European Commission (Horizon 2020 grant “Lifebrain”, 732592)), **MK** (UK MRC (MR/K013351/1, MR/R024227/1, MR/S011676/1), National Institute on Aging (NIH), US (R01AG056477), NordForsk (75021), Academy of Finland (311492), Helsinki Institute of Life Science Fellowship (H970)), **LG** (Monument Trust Discovery Award from Parkinson’s UK (J-1403) and the MRC Dementias Platform UK (MR/L023784/2)), **ASM** (NIH (R01AG056477, R01AG062553)). The views expressed are those of the authors and not necessarily those of the NHS, the NIHR or the Department of Health.

## Acknowledgements

We thank all the participating civil service departments; the British Occupational Health and Safety Agency; the British Council of Civil Service Unions; all participating civil servants in the Whitehall II Study; and all members of the Whitehall II Study team at University College London who so helpfully collaborated with us. The Whitehall II Study team comprises research scientists, statisticians, study coordinators, nurses, data managers, administrative assistants, and data entry staff, who make the study possible. Staff at the Wellcome Centre for Integrative Neuroimaging in Oxford, in particular research radiographers Michael Sanders, MSc, Jon Campbell, MMRTech, BcAppSc, Caroline Young, DCR(R), David Parker, BSc(Hons), who acquired the scans. Martin R. Turner, MA, MBBS, PhD, FRCP (Wellcome Centre for Integrative Neuroimaging, Oxford, United Kingdom), and his colleagues advised on incidental findings and taking over clinical responsibility for such participants. We are grateful to Professor Michael A. Chappell (DPhil) and Dr Thomas W. Okell (DPhil) for their advice on the FSL-BASIL tool for perfusion MRI analysis.

No compensation was provided for staff contributions to this study.

## Conflicts of Interest

Professor John E Deanfield reports provision of medical consulting for the Brain Protection Company Ltd. The remaining authors declare no conflicts of interest.

## Notes

### Clinical Trial

NCT03335696

### Author Declarations

University of Oxford Medical Sciences Interdivisional Research Ethics Committee

